# The current landscape of lifestyle and other non-pharmaceutical interventions in oncology in Africa: A review of registered clinical trials

**DOI:** 10.1101/2025.01.26.25321157

**Authors:** Monica Wangari, Eunice Njoroge, Grace Mburu, Brenda Mogeni, Luke Ouma

## Abstract

Cancer is a significant public health issue in low—and middle-income countries, especially in Africa. While lifestyle and/or behavioral interventions such as exercise and dietary modifications are known to improve patient outcomes and quality of life, critical evidence from resource-limited settings like Africa is scanty. Here, we present a review of clinical trials on the current landscape of cancer associated lifestyle interventions in oncology in Africa. We comprehensively reviewed non-pharmacological/ lifestyle cancer-related clinical trials in Africa on ClinicalTrials.gov and ICTRP trial registries from 01 July 2005 to 30 October 2024. We employed descriptive analyses, primarily frequency and percentage, to analyze and report the data. Overall, 53 trials matching the criteria were identified. Most of the trials were in Egypt (38/53, 97.4%), Kenya (4/53,10.3%), and Nigeria (5/53, 12.9%). The top cancer types for the trials in Africa were breast 24/62, 45.3%), colorectal (6/62, 11.3%) and acute lymphoblastic leukemia (4/62, 7.5%). Concerning sponsorship, most trials were universities/academia-sponsored trials (45/53, 84.9%), locally sponsored (37/53, 69.8%), conducted among patients (88.7%), and 84.9% cancer survivors. Most trials involved physical activity interventions (25/53, 47.2%) and psychological and psychological interventions (10/53, 18.9%), with (32/53, 60.4%) trials completed and (13/53, 24.5%) trials ongoing. Non-pharmacological interventions for the management of cancer appear to be nascent in Africa. Oncology trials are needed to ensure the effectiveness of non-pharmaceutical/lifestyle interventions in Africa, especially sub-Saharan Africa (SSA). In the current era of precision medicine, pharmacological, and lifestyle interventions, it no longer suffices to assume that interventions in high—and upper-middle-income countries will be effective in low-middle-income countries, especially Africa.

## 1. Introduction

Cancer is a major public health burden and the second leading cause of morbidity and mortality in Africa, with cancers of the cervix, breast, liver, colorectal and prostate ranked as the most common in the region [1,2]. By 2030, it is estimated that there will be a 70% increase in new cancer cases and approximately 1 million cancer-related deaths in the region annually [3]. This heavy disease burden is associated with ageing populations and adoption of unhealthy lifestyle habits, including increased physical inactivity, consumption of unhealthy diets, and harmful tobacco and alcohol consumption [3–7]. Besides, many countries in Africa are grappling with weak and inefficient health systems, delays in screening and treatment, unfamiliarity with cancer and related prevention strategies, and inaccessibility to quality and affordable care that further aggravate and contribute to higher mortality rates [1,8–10] Lifestyle interventions, such as those that promote a healthful diet and an active and non-smoking lifestyle, have demonstrated benefits in reducing adverse cancer-related outcomes and improving quality of life [11]. Lifestyle interventions are defined as having at least one additional component, including behavioural therapy, counselling, and stress management, in addition to both nutritional and physical activity components [12]. According to the World Health Organisation (WHO), nearly one-third of cancer-related mortality is preventable through lifestyle interventions and behavioural modifications, indicating that effective, evidence-based intervention strategies are urgently needed in Africa [13]. Today, several reviews have been conducted demonstrating that healthful lifestyle practices are preventative of the onset, progression, or recurrence of many cancers in both survivors and healthy individuals [14–17]. However, a critical analysis of the evidence shows these have largely been generated from high-income settings.

Current approaches to reducing the cancer burden in Africa have often faltered due to a lack of emphasis on lifestyle changes, revealing gaps in understanding and implementing preventive and management measures [18,19]. Given the cost and infrastructure challenges associated with precision oncology, lifestyle interventions could offer a viable solution for cancer prevention and management in Africa [20]. Despite substantial evidence of the role of healthy lifestyle habits in cancer prevention and management, the integration of lifestyle intervention in the African cancer landscape remains unexplored [21]. We need to understand the extent to which we leverage these low-cost but highly promising interventions in Africa.

This review synthesizes the current landscape of lifestyle interventions studied in interventional trials for the management of any cancer in Africa by analyzing clinical trial registry data.

## 2. Materials and Methods

### 2.1. Search methods

We conducted a search of registered trials on Clinical Trials.gov as well as the International Clinical Trials Registry Platform (ICTRP) from July 01, 2005, to October 30, 2024. The search utilized the following keywords: “cancer” OR “oncology” OR “lifestyle intervention” OR “non-pharmaceutical intervention”. A comprehensive list of search terms used in the trial registries developed for the PubMed database was subsequently adapted to each trial registry

(See S1 Appendix). The search was restricted to countries within Africa.

### 2.2. Eligibility criteria

To meet the inclusion criteria, trials must have been (1) *population:* cancer-specific and conducted in at least one African country; (2) *Intervention:* included lifestyle interventions (i.e. nonpharmacological intervention including a variety of diet or dietary interventions, physical activity, and behavioural components); (3) *Studies:* was an interventional study (i.e. non-observational, either randomised or non-randomised). We excluded studies where the primary intervention was pharmacological or that intervention was not aimed at supporting cancer patients. First, we assessed the relevance for the country, thereafter, the relevance of a trial to oncology, and the type of intervention. Studies were limited to prospective trials designed to evaluate non-pharmacological interventions registered on or after 1^st^ July 2005, which coincides with the mandatory registration of clinical trials [22].

### 2.3. Screening and data extraction

Three reviewers (M.W, G.M, and E.N) independently screened the trials identified by the searches, with a fourth reviewer (B.M) screening 10%, and using a pre-specified Microsoft Excel spreadsheet, data was extracted electronically by (B.M. and M.W). Reviewers were blinded to each other’s decisions throughout the screening process, and any conflicts identified from the screening were resolved with a fifth reviewer (L.O). The following information was extracted from the trials: trial identifiers, information source, country, cancer type, a summary of the study, sponsors, study type, study start date and end date, population, intervention, sex, and age eligibility. Discrepancies were resolved through ongoing team discussion. A summary of all information extracted from the trial registries can be found in S1 Table.

### 2.4. Data analysis and synthesis

All data visualisation and statistical analyses were performed using R software [23]. Descriptive statistics, mainly frequency and percentage, were used to describe and summarise the characteristics of the eligible trials. The findings are reported in a summary table and further described narratively.

## 3. Results

### 3.1. Trial design characteristics

The initial search of the trial registries identified 225,294 trial records. Of these, 7725 trial records met the eligibility criteria of a study conducted in at least one African country and were further screened for eligibility. A total of 53 studies across nine African countries met our inclusion criteria. A complete list of trial records included in this review is provided in the S2 Table.

Table 1 provides a summary of study characteristics for relevant studies evaluating lifestyle and other non-pharmaceutical interventions in Africa. Most trials in our review were randomised (92.5%) and blinded studies (71.7%), were ongoing (32/53) or completed (13/53), recruited patients as the primary beneficiary or recipient of the intervention (88.7%), and were in adult populations (69.8%). Notably, these cancer studies were not limited to cancer patients – about six studies investigated interventions in carers (11.3%), and a further eight studies sought to understand the efficacy or effectiveness in cancer survivors (15.1%). Furthermore, we also observed a growing interest in studies recruiting paediatric populations. Nearly one-third of lifestyle interventions in oncology trials included paediatric populations (30.5%).

**Table 1:** Summary characteristics of relevant studies included in the review.

| Characteristic |  | Sub-Saharan African country |  | Total (N=53) |
| --- | --- | --- | --- | --- |
|  |  | NO (N=39) | YES (N=14) |  |
| Completion status | Completed | 25 (64.1%) | 7 (50%) | 32 (60.4%) |
|  | Ongoing | 11 (28.2%) | 2 (14.3%) | 13 (24.5%) |
|  | Unknown | 3 (7.7%) | 3 (21.4%) | 6 (11.3%) |
|  | Withdrawn | 0 (0%) | 2 (14.3%) | 2 (3.8%) |
| Intervention population | Carer-nurse, relative | 2 (5.1%) | 3 (21.4%) | 5 (9.4%) |
|  | Patient | 37 (94.9%) | 10 (71.4%) | 47 (88.7%) |
|  | Carer & patient | 0 (0%) | 1 (7.1%) | 1 (1.9%) |
| Post-surgical intervention | No | 22 (56.4%) | 12 (85.7%) | 34 (64.2%) |
|  | Yes | 17 (43.6%) | 2 (14.3%) | 19 (35.8%) |
| Cancer survivors' study | Yes | 5 (12.8%) | 3 (21.4%) | 8 (15.1%) |
|  | No | 34 (87.2%) | 11 (78.6%) | 45 (84.9%) |
| Study sponsor | Academia/university | 32 (82.1%) | 13 (92.9%) | 45 (84.9%) |
|  | Hospital | 4 (10.3%) | 1 (7.1%) | 5 (9.4%) |
|  | Investigator-led study | 2 (5.1%) | 0 (0%) | 2 (3.8%) |
|  | University Hospital | 1 (2.6%) | 0 (0%) | 1 (1.9%) |
| Local study sponsor | No | 3 (7.7%) | 13 (92.9%) | 16 (30.2%) |
|  | Yes | 36 (92.3%) | 1 (7.1%) | 37 (69.8%) |
| Sex | All | 21 (53.8%) | 7 (50%) | 28 (52.8%) |
|  | Female | 17 (43.6%) | 7 (50%) | 24 (45.3%) |
|  | Male | 1 (2.6%) | 0 (0%) | 1 (1.9%) |
| Study population | Adult | 26 (66.7%) | 11 (78.6%) | 37 (69.8%) |
|  | Child | 8 (20.5%) | 0 (0%) | 8 (15.1%) |
|  | Child & Adult | 5 (12.8%) | 3 (21.4%) | 8 (15.1%) |
| Paediatric trial | False | 25 (64.1%) | 8 (57.1%) | 33 (62.3%) |
|  | True | 13 (33.3%) | 3 (21.4%) | 16 (30.2%) |
| Randomised trial | No | 3 (7.7%) | 1 (7.1%) | 4 (7.5%) |
|  | Yes | 36 (92.3%) | 13 (92.9%) | 49 (92.5%) |
| Masking/blinding | Double | 8 (20.5%) | 2 (14.3%) | 10 (18.9%) |
|  | None | 11 (28.2%) | 4 (28.6%) | 15 (28.3%) |
|  | Single | 20 (51.3%) | 8 (57.1%) | 28 (52.8%) |

Regarding gender distribution, Females were the single most included gender, although over half of the studies (52.8%) included both sexes. Only one study exclusively recruited male study participants (NCT06605846). This is particularly unsurprising, as this is largely determined by the cancers under investigation—a higher number of breast cancer studies implied a higher number of female-only studies.

Most studies in this review were locally sponsored studies (69.8%). However, studies in SSA were far more commonly locally sponsored studies compared to those outside of SSA. For instance, 92.9% of studies in this review from SSA were funded by external sponsors, while 92.3% of studies outside SSA were funded by local sponsors. For both studies in and outside SSA, universities/academia were the major sponsors of studies included in the review (84.9%). For SSA studies, these were academic institutions outside of the continent. Additionally, post-surgical interventions – studies evaluating the effectiveness of an intervention following a surgical procedure – were fairly common. About one-third of studies (35.8%) evaluated post-surgical interventions.

As reported in Table 1, we notice differences in nearly all the characteristics across studies within and outside SSA. In **Fig 1**, we summarise a few quantitative summary characteristics of studies.

**Fig 1.**
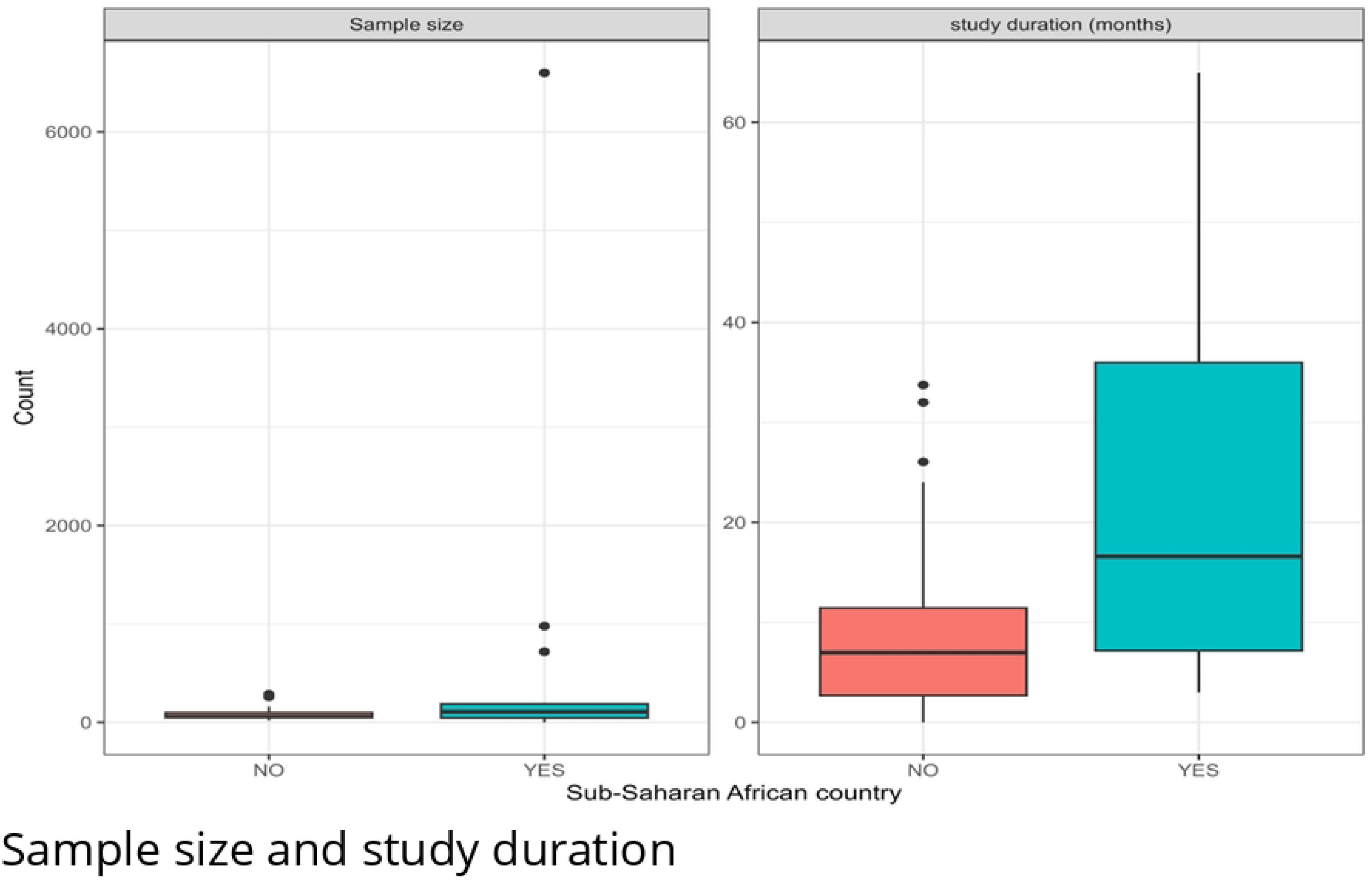
Sample size and study duration of oncology studies in Africa.

Studies in SSA were, on average, larger by sample size (median: 109 vs. 60 patients) and conducted for a longer duration (median: 16.6 months vs. 7 months) compared to those outside of SSA.

### 3.2. Trends in lifestyle intervention studies in oncology

There has been a steady increase in studies investigating non-pharmaceutical and lifestyle interventions in Africa. The earliest study in our review was initiated in 2009 (ACTRN12612000256875), and since then, there have been, on average, 2-3 new registrations per year until 2022, from which there have been 5+ new trials per year. The highest record of trial registrations was observed in the years post-pandemic (2022 onwards), especially in 2023, where 16+ studies were registered.

The study trends reported in **Fig 2** demonstrate geographical disparities in study coverage across the continent (**Fig 2A)**

**Fig 2:**
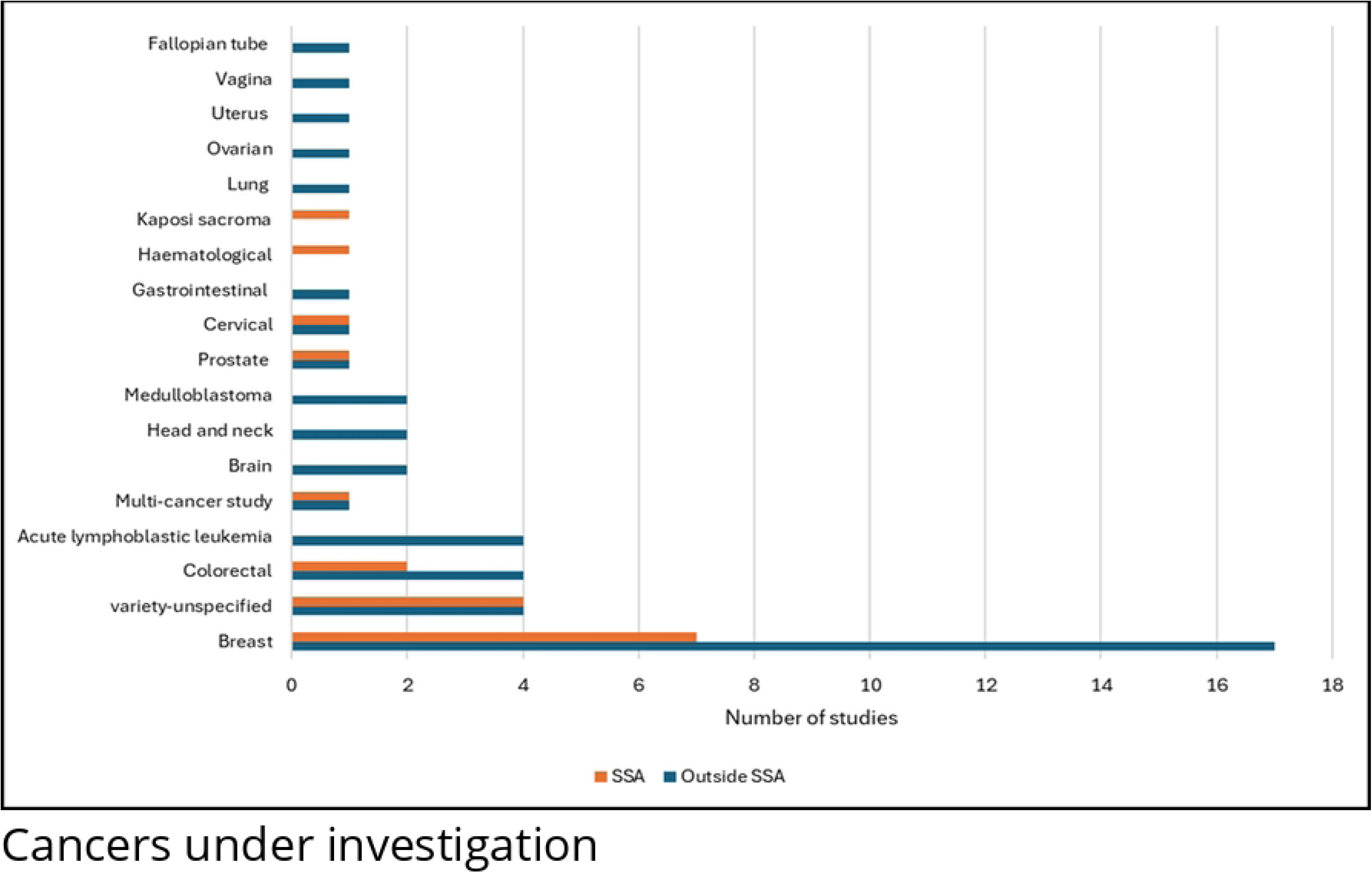
Trends in oncology non-pharmaceutical intervention studies. **Fig 2A: Trends in trial registration across regions of Africa; Fig 2B: Trends in trial registration across sub-Saharan Africa**

Firstly, we observe that the majority of trials (73.5%) were conducted outside of Sub-Saharan Africa **(Fig 2B).** The trends show consistently growing numbers of oncology lifestyle intervention studies ongoing outside of SSA, especially in Northern Africa. Besides the North, the East African region hosted a considerable number of studies, followed by West Africa. There were only two studies from Southern Africa, one initiated in 2016 and another in 2023.

### 3.3. Types of cancers under investigation

**Fig 3** summarises the characteristics of the types of cancers under investigation in the included trials. Our findings demonstrate that most trials were interventions in breast cancer (n=24, 45.3%), followed by colorectal (n=6, 11.3%), while a small fraction (n=8, 15.1%) did not specify the cancer types. Overall, the most prevalent cancer in SSA included breast cancer (50%), colorectal (14.3%), cervical, prostate, haematological, Kaposi sarcoma each (7.1%), and variety-unspecified cancer (28.6%). A fraction of the trials targeted multiple cancers (n=2, 3.8%).

**Fig 3:**
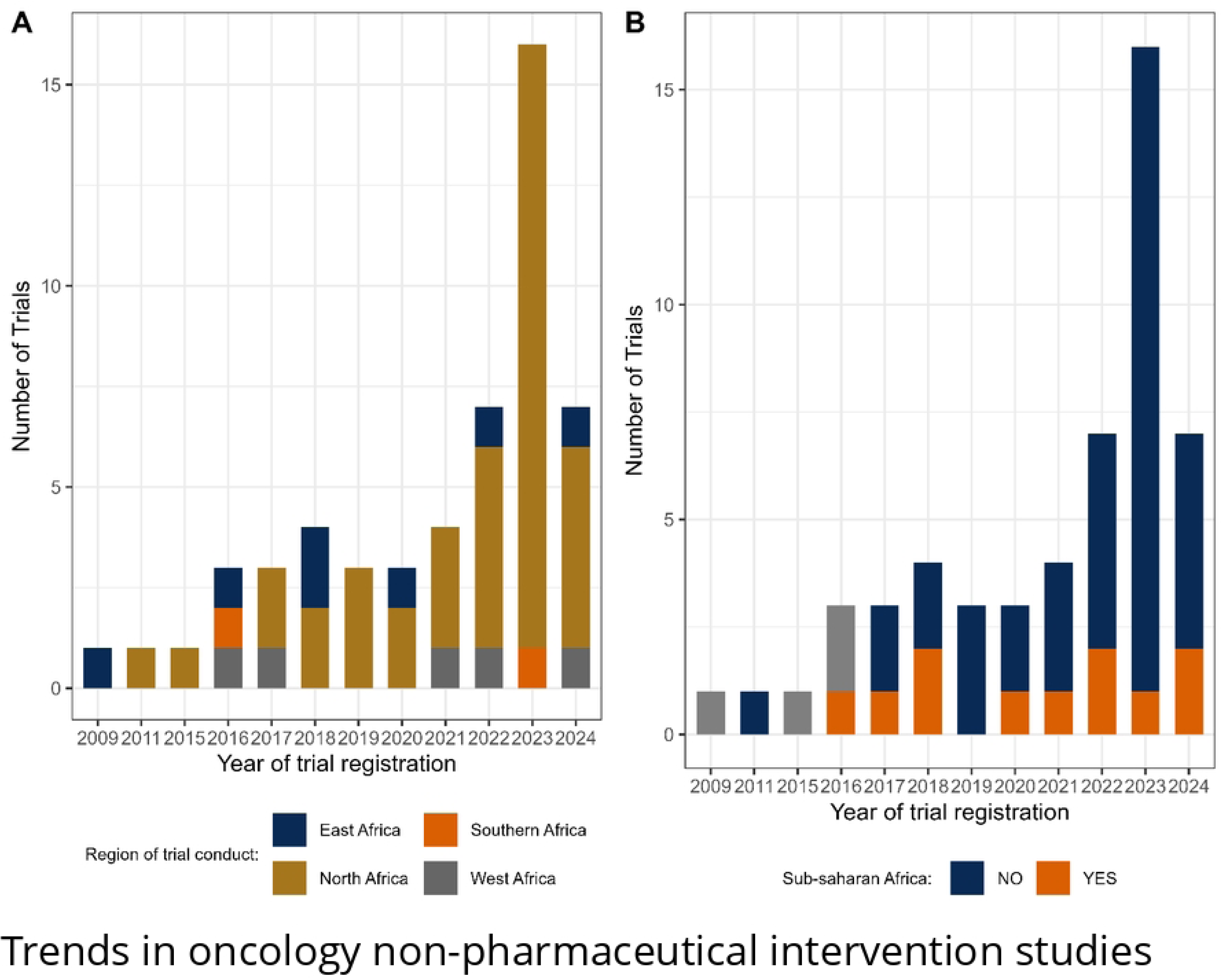
Cancers under investigation for lifestyle and other non-pharmaceutical interventions in Africa.

### 3.4. Types of lifestyle interventions under investigation

We present a summary of the intervention under investigation across various cancers in Table 2. A total of 64 lifestyle interventions were identified across six broad categories: dietary, educational, financial, physical activity, psychological and technological interventions. Physical activity interventions were the most prevalent, constituting (43.8%, n=28) of all the interventions, followed by psychological intervention (23.4%, n=15) and dietary interventions (9.4%). Of the two leading interventions, only 28.6% (n=4) of each were SSA studies. Educational, financial, and technological interventions were less frequently reported, accounting for 7.8% (n=5), 6.3% (n=4), and 9.4% (n=6) of interventions, respectively. A notable number of cancer trials involved more than one modality of intervention.

**Table 2:** Summary of interventions under investigation across various cancers.

| Conditions | Dietary intervention | Educational intervention | Financial intervention | Physical activity | Psychological intervention | Technological intervention | Total |
| --- | --- | --- | --- | --- | --- | --- | --- |
| Acute lymphoblastic leukaemia | 1 | 0 | 0 | 3 | 0 | 0 | 4 |
| Breast | 2 | 0 | 1 | 12 | 6 | 2 | 23 |
| Colorectal | 2 | 1 | 1 | 1 | 1 | 0 | 6 |
| Gastrointestinal | 1 | 0 | 0 | 0 | 0 | 0 | 1 |
| Variety-unspecified | 0 | 4 | 1 | 0 | 2 | 2 | 9 |
| Lung | 0 | 0 | 0 | 1 | 0 | 0 | 1 |
| Brain | 0 | 0 | 0 | 2 | 0 | 0 | 2 |
| Breast | 0 | 0 | 0 | 3 | 0 | 0 | 3 |
| Head and neck | 0 | 0 | 0 | 2 | 0 | 1 | 3 |
| Kaposi sarcoma | 0 | 0 | 0 | 1 | 0 | 0 | 1 |
| Medulloblastoma | 0 | 0 | 0 | 2 | 0 | 0 | 2 |
| Prostate | 0 | 0 | 1 | 1 | 0 | 0 | 2 |
| Haematological | 0 | 0 | 0 | 0 | 1 | 0 | 1 |
| Cervical | 0 | 0 | 0 | 0 | 1 | 1 | 2 |
| Ovarian | 0 | 0 | 0 | 0 | 1 | 0 | 1 |
| Uterus | 0 | 0 | 0 | 0 | 1 | 0 | 1 |
| Vagina | 0 | 0 | 0 | 0 | 1 | 0 | 1 |
| Fallopian tube | 0 | 0 | 0 | 0 | 1 | 0 | 1 |
| Totals | 6 | 5 | 4 | 28 | 15 | 6 |  |

Physical activity and psychological intervention were frequently applied interventions to breast cancer, accounting for 52.8% and 26.1%, respectively, while dietary interventions were limited but utilised mostly in breast and colorectal cancer. Furthermore, educational and technological interventions were primarily used in various unspecified breast cancers.

### 3.5. Geographical distribution

**Fig 4** and S3 Table present the geographical distribution of the trials included in this review. Of the 53 studies included in this review, the vast majority (98.1%, n=52) were single-country studies, with one conducted across multiple countries and located within SSA. Thirteen of the 52 studies were conducted exclusively in SSA.

**Fig 4:**
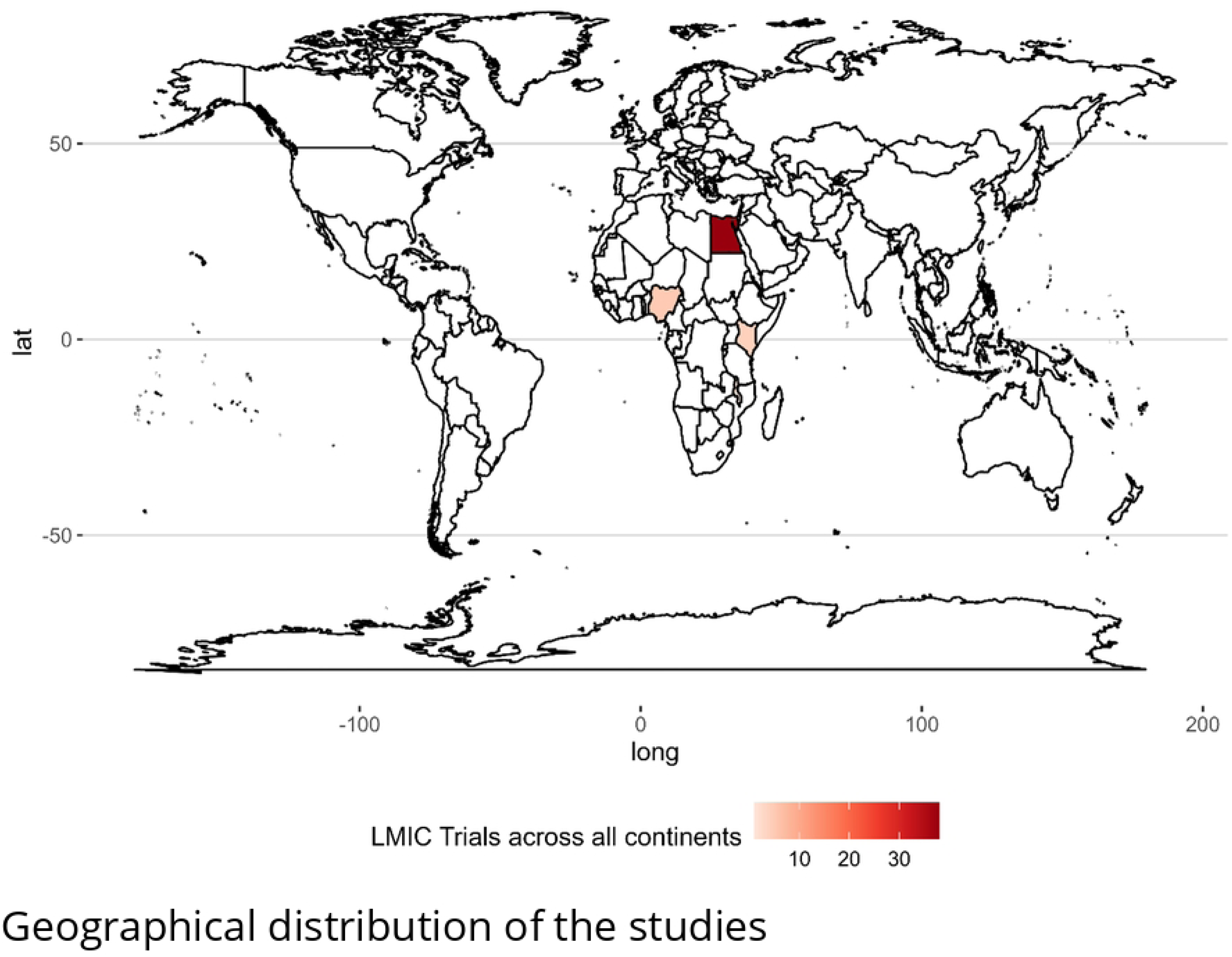
Geographical distribution of the studies.

Regionally, more than half of these trials (39/53, 73.6%) were conducted in North Africa, followed by 13.2% (n=7) in East Africa, 9.4% (n=5) in West Africa and 3.8% (n=2) in southern Africa. Notably, there were no trials focused on Central Africa. A strong research concentration was found to have taken place in Egypt (n=38, 97.4%), accounting for nearly all North African studies. In SSA, studies were more evenly distributed across Kenya and Nigeria, with four studies each (10.3%). Ethiopia, Malawi, Rwanda, Uganda, Tunisia, and Botswana each contributed (2.6% per country), demonstrating a relatively fragmented research landscape in these regions.

## 4. Discussion

As a leading cause of mortality globally and in Africa, innovations for the treatment and management of cancer are of interest to the research community. In resource-poor settings such as Africa, affordable, cost-effective, and impactful public health interventions are much needed to counter the ever-growing cancer threat and the burden it exerts on already weak health systems [24,25]. Whilst oncology research in Africa continues to gather pace, this review presents the current status quo of non-pharmaceutical and lifestyle interventions for cancer management in Africa. Clinical trials of non-pharmaceutical interventions offer valuable insights into the landscape of cancer research in Africa, specifically focusing on lifestyle interventions for cancer management.

Our review demonstrates that similar to pharmaceutical interventional studies, there is a dearth of studies focusing on lifestyle interventions in oncology in Africa and largely much of sub-Saharan Africa (SSA). Studies were generally smaller in a few classes of cancers and spread in a few countries. Reviews that have largely focused on pharmaceutical interventions have reached similar conclusions – the pace of cancer research in Africa is lagging [1,26]. Of the trials ever initiated, ongoing, withdrawn or completed, we observed significant geographical disparities in the distribution of trials– the dominance of North Africa, few studies in SSA, sparse representation from southern Africa, and no trials reported in central Africa. The uneven distribution of trials across Africa reflects existing regional differences in research infrastructure, funding availability [27] accessibility to high quality care [26] and disease burden prioritisation [28,29]. For instance, it was unsurprising to see that studies outside SSA were more commonly locally sponsored, while studies within SSA were often funded by sponsors outside the region. Furthermore, the scant data from central Africa indicates the lag in the clinical research community’s responsiveness to emerging health challenges. Such health emergencies may drive the prioritisation of research efforts and facilitate rapid advancements in the understanding and management of infectious diseases [30].

Although a majority of studies focused on adult cancer patients, there is a discernible shift towards recognising the importance of diverse beneficiaries, as evidenced by the marginal but growing inclusivity of paediatric populations, caregivers, and cancer survivors. These trends reflect an evolving understanding of the multifaceted needs within this field, extending beyond merely addressing the requirements of active cancer patients. Still, female participants dominate most studies, highlighting the dominance of studies particularly in female breast and gynaecological cancers (over half of the studies), and there is a pressing need to expand the scope for male inclusivity in trials. This is of importance given the higher prevalence of certain male cancers, such as prostate, across the continent.

Our analysis of the landscape of interventional studies targeting cancer shows that physical activity and psychological interventions are the predominant modalities. However, the representation of these interventions within SSA was small, about one in three studies. We see a significant gap in evidence-based interventions where resources for cancer management are constrained particularly in SSA. Furthermore, educational, financial, and technological interventions are markedly underrepresented, presenting a unique opportunity for future research. A contextual understanding of these research priorities is essential for developing a comprehensive and culturally relevant cancer management strategy that aligns with the known needs and challenges faced by diverse populations in SSA.

The evaluation of the funding landscape for clinical research in Africa reveals a predominant reliance on local support, particularly from academic institutions/universities, which suggests a limited influence of commercial entities within the realm of clinical research in the continent. However, SSA was more dependent on external funding for nearly all the studies, reflecting challenges in securing local research funding. This reliance on international sponsors emphasises the need to strengthen local research funding mechanisms to enhance the sustainability of research in SSA. While industry sponsorship is a key driver for advancing oncology drug development, public and academic institutions significantly contribute to funding research that tackles important health issues, such as public health interventions, which may be less equitable but are equally critical.

The review by Odedina et al. [26] shows that Egypt and South Africa are the leading countries where pharmaceutical companies have supported clinical trials. On the contrary, there were no industry partners funding studies in this review. It is, therefore, evident that industry support is exclusively focused on pharmacological interventions. Even then, the shift towards precision oncology in high-income countries has not yet translated effectively to Africa, where access to innovative targeted therapies remains limited [31,32]. Several targeted oncology therapies have shown significant promise in improving patient outcomes, yet access to some of these life-saving therapies remains limited in regions such as Africa. Indeed, these raise the question and need for the adoption of effective, scalable, affordable, and high-impact public health interventions.

There are a few limitations to this study. First, we rely exclusively on clinical trial evidence from registry data. However, by reviewing data from trial registries we obtain a clearer picture of the status quo, identifying both that would otherwise not appear in research publication databases. Secondly, we did not focus on any non-interventional studies which may otherwise provide additional insights into the status quo.

## 5. Conclusions

This review mapped the landscape of lifestyle and other non-pharmaceutical interventions in oncology. Although the benefits of lifestyle interventions across several cancers have been characterized elsewhere, research from Africa is scanty and largely domiciled outside of SSA, where there is less burden compared to SSA. Our findings suggest that the full spectrum of lifestyle intervention-induced health outcomes is yet to be carefully explored for most, if not all, cancers in Africa. Furthermore, given the lag in precision medicine research in Africa and the high associated cost of promising targeted therapies, there is a greater need for research prioritization, given the affordability, scalability, and access to promising lifestyle interventions.

## Data Availability

All relevant data are within the manuscript and its Supporting Information files.

## Supporting Information

**S1 Appendix. Literature search strategy.**

**S1 Table. Data extraction template.**

**S2 Table. List of included studies.**

**S3 Table. Supplementary results, geographical distribution.**

